# A rapid systematic review and case study on test, contact tracing, testing, and isolation policies for Covid-19 prevention and control

**DOI:** 10.1101/2020.06.04.20122614

**Authors:** Sheng-Chia Chung, Sushila Marlow, Nicholas Tobias, Ivano Alogna, Alessio Alogna, San-Lin You

## Abstract

**Objectives:** To conduct a rapid review on the efficacy and policy of contact tracing, testing, and isolation (TTI) in Covid-19 prevention and control, including a case study for their delivery.

**Method:** Research articles and reviews on the use of contact tracing, testing, self-isolation and quarantine for Covid-19 management published in English within 1 year (2019 to 28^th^ May, 2020) were eligible to the review. We searched MEDLINE (PubMed), Cochrane Library, SCOPUS and JSTOR with search terms included “contact tracing” or “testing” or “self-isolation” or “quarantine” in the title in combination with “Covid-19” or “COVID-19” or “coronavirus” in the title or abstract. Studies not associated with TTI or Covid-19 or being solely commentary were excluded. A narrative synthesis with a tabulation system was used to analyse studies for their diverse research designs, methods, and implications. Data for the case study were obtained from the Ministry of Health and Welfare and Centers for Disease Control Taiwan.

**Results:** Among the 160 initial publications, 48 eligible studies are included in the review. Included studies applied various designs: experiments, clinical studies, Government Documents, systematic reviews, observational studies, surveys, practice guidelines, technical reports. A case study on TTI delivery is summarised based on policy and procedures in Taiwan.

**Conclusions:** The information included in the review may inform the TTI program in the UK.

## INTRODUCTION

Since the initial Covid-19 cases appeared in November 2019, the pandemic has led to 7,085,894 new cases worldwide and 405,170 deaths.(1) In the UK, as of 8^th^ June, 2020, there have been 287,399 cases of COVID- 19, including 40,597 deaths.(2) To stop the transmission of the disease, non-essential activities have been restricted in the country since 21^st^ March. The strategy has effectively lessened the spreading of the disease, and in May, there has been a steady decline in Covid-19 incidence and mortality.

To control Covid-19 in the second phase of the pandemic, the UK government launched the National Health System Test and Trace program on 27^th^ May. Contact tracing, testing, self-isolation and quarantine measures are commonly used to contain infectious diseases, and countries who have implemented these strategies effectively have reported low to no Covid-19 endemic. However, there are controversy on the legislative, ethical, and human right aspects of the delivery of tracing, testing and isolation (TTI). For example, there have been concerns on personal data protection, privacy, and oversight in contact tracing. Potential unintended consequences (such as discrimination and exclusion) and conflict of interests may also put a stain on the public trust, which is crucial in a collective pandemic response.

Since the first Covid-19 case reported in Taiwan on 21^st^ January 2020, the country has implemented different TTI measures and ended the urgent Covid-19 on the 7^th^ June. Taiwan’s experience may inform the pragmatic aspects in the delivery of TTI. The objective of the rapid review is to collate and interpret the existing evidence on the efficacy of TTI programs and policies in their delivery. A case study of TTI delivery in Taiwan is included in the report. Results of the review may inform towards an effective and benevolent strategy for test, trace and isolate in the UK.

## METHODS

### Systematic review

#### Eligible studies

are research articles and reviews evaluating the effectiveness of contact tracing, testing, self-isolation and quarantine on Covid-19 management published in English within 1 year (May, 2019 to May, 2020).

#### Search

We searched MEDLINE (PubMed), Cochrane Library, SCOPUS and JSTOR. The search terms included “contact tracing” or “testing” or “self-isolation” or “quarantine” in the title in combination of “Covid-19” or “COVID-19” or “coronavirus” in the title or text. The date last searched is 28^th^ May 2020. The subject of the study was limited to human. Relevant reports and literature from the reference of the articles or suggested to the review team were included.

#### Study selection

In MEDLINE search, we selected clinical studies, clinical trials, evaluation studies, government Documents, journal articles, meta-analyses, reviews, systematic reviews, multicentre studies, observational studies, practice guidelines, pragmatic clinical trials, technical reports. We selected studies whose subject area were in medicine or social sciences in SCOPUS, and research reports in JSTOR (excluding book chapters). We used title, abstract and content screening to remove studies not related to the TTI or Covdi-19.

We used a standardised list of questions to synthesise information from the eligible studies in the review (supplementary session one). It collected background information on study design, method, results, main findings, and limitations according to PICOS together with the list of policy review questions. Summary measures of each participant study, when available, were assessed and the risk of bias in the results and limitation of participant studies evaluated.

Information for the case study was obtained from the Centers for Disease Control Taiwan (https://www.cdc.gov.tw/en/Disease/Sublndex/ together with a detailed documentation of the crucial policy milestone for covid-19 response in Taiwan from the ministry of health and welfare website available from 7^th^ June 2020: https://covid19.mohw.gov.tw/en/mp-206.html.

## RESULTS

We identified 60 studies from PUBMED (Medline), 43 studies from SCOPUS, 56 from JSTOR and 1 from Cochrane library and 4 articles from additional sources. Among the 160 initial publications, we removed 30 duplicate studies, 81 items that were not directly focused on Covid-19 or strategy of testing, contact tracing or isolation, or not in a research report format (such as editorial, audio interview).

### Overview of TTI

In the rapid literature review, Nussbaumer-Streit B et al. summarised a combination of quarantine and other nonpharmaceutical interventions can significantly reduce Covid-19 incidence and mortality.(3) From modelling studies, Manchein et al suggested the growth of Covid-19 cases following a power-law pattern than exponential, indicating the propagation of virus may be subject to free networks, fractal kinetics and show small world features. Their modelling emphasised the importance of social distancing to slow the spread of Covid-19, in concordance with the modelling findings by Sjödin et al.(4) A potential limitation of the modelling may be that limited testing can underestimate the true number and growth rate of Covid-19 cases, as showed by Omari et al.(5) Testing can facilitate case finding and support social distancing by identifying asymptomatic cases.(6) Chen et al. described elements to effectively contain SARS-Cov2 transmission include quarantine, isolation, and surveillance of disease progression after contact tracing.(7) An effective TTI and Covid-19 response requires the collaboration of the population as a whole, while it can leverage technology and data integration for effectiveness.

#### 1. How would a 24 hr turn-around from sampling to result actually work?

Lagier et al. reported the procedure of SARS-Cov2 RT-PCR testing for repatriated French citizens.(8) Samples from 337 passengers on the repatriation flights were tested. The time from sample-to-results was 4 h and 50 min (290 min) and 6 h and forty (400 min) for the samples from the first returning flight. The laboratory thus optimised the testing strategy by prioritising the extraction of RNA from the samples and reduced the time from sample to results to 2 h and 50–55 minutes (170–175 minutes).

SARSCoV-2 RNA could be detected in stool, blood, or urine if undetected in upper respiratory tract specimens.(9) The acceleration of RNA testing has also been promoted by Yan et al who suggested that a key action would be place samples in reagents containing guanidine salts, for example TRIZOL, TRIZOL LS, or AVL buffer to inactivate the virus and protect RNA. Pooling of samples, using statistical algorithms was suggested by Abdalhamid et al to reduce the number of tests required and expedite the testing process.(10) The optimal pool sizes were selected based on the prevalence rate. A similar three-stage pooling system was reported by Eberhardt et al to have a 3.8 improvement factor at 12% prevalence.(11)

##### 1–1. infrastructure and procedure of a sampling-to-results process that is 24 hours or less

Hill et al. reported the high efficiency of drive-through SARS-Cov2 testing from the experience of NHS Lothian(12) with patients being informed of their results within 24 to 36 hours. However, this service was only applicable to individuals with access to a private vehicle and fit to drive.

Alternately, Binniker has outlined a system of setting up testing centres in all available clinics, as deemed safe and trustworthy by a responsible body.(13) These can be used to promote the role of local communities in testing and encourage people to take tests. Similarly, Gupta et al have described the response of laboratories in universities and clinics in India being converted to testing centres.(14) Tolia et al confirmed the feasibility of an in-house Covid-19 testing with the clinical laboratory of a hospital.(15)

##### 1–2. Testing strategy and testing during emergency

The case definition of Covid-19 changed frequently and quickly in the initial stage of the epidemic, as the knowledge about the disease increased with time. Change in case definition usually followed by a rise in the number of new cases reported and testing required for suspected cases. In the early stage of the epidemic, there were limited test kits and qualified laboratories, and testing was prioritised for suspected cases.(16) The definition of suspected cases differed from country to country, depending on the progression of the epidemic. A broader criteria for case definition is used for countries primarily concerned about imported cases, or when community transmission is possible. Restricting testing to persons with relevant travel, contact or work history may leave undetected individuals whose source of infection cannot be identified.

To investigate the potential SARS-Cov-2 local transmission, the Dutch National Outbreak Management Team conducted a rapid screening among health care workers in an area of suspected Covid-19 outbreak. Testing was done followed the national protocol and carried out either locally or in central laboratories. Nine hospitals were requested to sample health care workers on Saturday, 7^th^ March, and testing results were due on Monday, 9^th^ March. The SARS-Cov-2 positive rate was 4.1% among 1097 tested; the local outbreak was confirmed, and the government immediately rolled out contingency plan.(17)

##### 1–3. minimising false-positive and false-negative results

In a review conducted by Younes et al, the RT-PCR testing kits developed in the US, France and Germany had a high sensitivity of 95% but specificity was not published.(18) A negative test does not rule out the disease, as in practise individuals with initial RT-PCR negative can become positive with repeated tests.(19) The sensitivity may have reduced, as a result of trace amount of virus sampled according to the sampling site and viral shedding at the time of sampling.(19, 20) To reduce false negative results, a multiple-sampling diagnosis can be applied in the testing protocol (case study session 1–3) or based on the clinical judgement. Point of care testing can be utilised for large-scale testing during emergencies. However, Döhla et al reported low sensitivity of point of care testing (36.4%) compared to RT-PCR in a small group of 39 Covid-19 patients.(20)

Yan et al have suggested that the human RNase P gene could be amplified as an internal control to reduce false-negative results and template volume could be increased to improve the detection sensitivity.(9) The test statistics as of March 2020 in India, as reported by Gupta et al, revealed 100% accuracy in negative tests while only 5 in 7 positive tests were verified as true.(14) The reconfirmation of positive tests were therefore outsourced to a centralised agency, namely; the National Institute of Virology (NIV), which reprocessed positive tests to quantify the accuracy of testing. In addition, the importance of continued testing of relevant markers in serious cases was stressed by Favaloro et al, arguing that more accurate markers could be found to indicate the presence of the disease or the degree of severity. Suggested markers are D-dimer and PT, APTT, fibrinogen and platelet counted to gauge the chances of a high risk patient.(21)

With the limitation in the validity of sampling and testing, the public should be made aware of potential false positive or false negative results. Especially in the case of negative test results, repeated testing is often required if symptoms persist.

##### 1–4. Linked data to facilitate active case finding

In order to identify cases from individuals at high risk of infection, such as those in contact with Covid-19 cases, information of the contacts can further linked to clinical data or claim data to track their health status.(7) Hospitalisation due to pneumonia can be identified for follow-up and SARS-CoV-2 testing. Screening for high-risk population (such as health care workers) has been used for active case finding.(22)

#### 2. What local governance and partnership structures are required, and what would a local “outbreak team” look like?

In Taiwan, local outbreak teams are responsible for contact tracing, quarantine enforcement and risk management.(23) Details on the sectors involved in outbreak control, their roles and responsibilities are summarised in the case study (session 2).

#### 7. Adherence to isolation and local support needs

##### Inbound traveller quarantine

Singapore imposed a 14-day “Stay Home Notice” (SHN) on visitors and returning residents from Covid-19 endemic areas since January 2020 and for all inbound traveller from 9^th^ April 2020. Upon arrival, travellers undergo a 14-day mandatory stay in government-designated hotels. During this period, they are not allowed to leave their individual rooms where specific infectious disease prevention procedure implemented.(24)

##### Timely identification of contacts

Ferretti et al. reported an estimated reproduction number of SARS-Cov2 of 2.0, of which 0.9 was infection occurred during the pre-symptomatic stage, suggesting about half cases were infected by Covid-19 patients before symptom onset.(25) It thus is essential to stop the transmission by instantaneously finding cases and their contacts for treatment, self-isolation or quarantine. However, the conventional epidemiological contact tracing, which relies on personal interviews is labor-intensive and time-consuming, and may not be feasible during large-scale epidemic.(7)

##### Digital tools to facilitate TTI

Telecommunication provider-based measures are more efficient than voluntary-based digital tools (such as Apps). Democratic nations adopt provider-based measures openly discuss its surveillance architectures, while less democratic nations tend not to or hide such information from the public.(26)

Types of telecommunication provider-based measures may include:

1. mapping the amount of anonymized cell phone movement in a particular area (Germany, Austria, Italy)
2. Base station triangulation to approximate cell phone location. (Taiwan)
3. Access the A-GPS data generated by the phone (Israel)

Voluntary provision of data including

1. App recording device within a contact range via Bluetooth technique. (Singapore, Austria),
2. App recording daily symptoms (South Korea, Taiwan, Poland),
3. QR code for entry or exit places (China).

In the Singaporean, for example, during the aforementioned stay home notice (SHN) period, text messages are sent at random times to the mobile phone of individuals undergoing SHN, and the receiver to report his or her GPS location via the weblink specified in the message. If the respond is not received within an hour, a video calls or house visits will be arranged by the health authority.(24)

Case studies for details in digital TTI tools in South Korea, Taiwan, Singapore, China, and Israel and how they are used in Covid-19 prevention and control can be accessed in the Appendix A of the report “Pandemic Mitigation in the Digital Age: Digital Epidemiological Measures to Combat the Coronavirus Pandemic.”(26)

##### Support for individuals to comply with self-isolation

Webster et al. identified the factors associated with adherence were the knowledge people had about the disease and quarantine procedure, social norms, perceived benefits of quarantine and perceived risk of the disease, as well as practical issues such as running out of supplies or the financial consequences of being out of work.(27) Self-isolation or quarantine may not be affordable for low-wage and gig workers.(28)

To address such financial needs, the Singaporean government provides self-employed persons and businesses with employees undergoing self-isolation or quarantine (SHN) a $100 per day support. Deliveries of food and supply can be arranged for individuals serving SHN, through a designated hotline. By law, landlords and dormitory operators cannot evict tenants serving SHN. For individuals where their residence may not be suitable for SHN, the government offers hotels as an alternative location.(24)

The psychological stress(29–31) or lack of physical activities(32) during self-isolation may have adversely effect on mental or physical health. In a rapid review, Brooks et al reported individuals in quarantine experiencing confusion, anger, post-traumatic stress symptoms, due to reasons such as fear of infection, frustration, boredom, and inadequate supplies.(30) Strategies to mitigate the adverse effects include timely and sufficient information to reduce uncertainty and facilitate cooperation, minimising quarantine interval to no longer than necessary, and providing adequate supplies for individuals serving self-isolation.

Razai et al. suggested validated psychological screening tools, such as Patient Health Questionnaire-4 for anxiety and depression and UCLA Loneliness Scale, can be utilised to identify individuals who need support. The care can be delivered by counselling, coaching via telephone or online video consultations.(31) Nonmedical social prescribing, such as visual choir, online theatre, performances or classes for exercise or art can reduce boredom. Communication by social media can ease loneliness. Banskota et al summarised smartphone apps to assist older adults coping with self-isolation, including apps for social networking, food and drinks delivery, medical consultation, health and fitness.(33) Meinert et al described an agile process to develop an app for older people and their families to improve well-being while observing social distancing rules.(34)

#### 4. How would real-time data management, linkage of datasets, and dashboards be developed, and who would “own this”

##### APP and GPS

In South Korean, the government uses several digital databases to facilitate contact tracing, such as electronic health records, phone-based GPS, card transaction records, and closed-circuit television.(35) Yasaka et al have proposed the use of an app with three guides, requiring users to create checkpoints (e.g. public spaces or shops with a QR code that can be scanned to the app), check risk level and report positive status.(36) Likewise, in China, QR code-based app are used for limiting the movement of suspected Covid-19 patients and displays a green, amber, or red code that is required for existing or entering places, and it remains unclear the digital surveillance architecture and data protection mechanisms for the personal data being collected and used.(37) Public scrutiny for data protection and privacy have been raised for both the Chinese and South Korean apps.

The efficacy of mobile positioning data (except in the case of 2G phones) was studied for use in Nigeria by Ekong et al. based on the systems used in South Korea, Singapore and China.(38) A legal framework for data protection was suggested when implementing these systems and a third-party agreement for data use.

##### Mobile geopositioning data

The mobile geopositioning method has been used in studying the mobility, disease connectivity, and health risk in travellers.(39) The mobile positioning measures are up to 150 meters from the actual location, and thus reduce the risk of undermining individual privacy.(7) Taiwan has applied a geopositioning method to facilitate the compliance of home isolation or quarantine (case study session 7.1) and rapid identifying contacts of suspected large-scale outbreak. An example is the identification of 627,386 contacts of the 3,000 Diamond Princess passengers touring in Taiwan during Covid-19 outbreak.(7) The process took a day and text messages for self-health management were sent to all contacts. Contact data were linked to healthcare administrative system for follow-up and testing of symptomatic contacts (Supplementary session three). The geopositioning method has lessened the pressure on health authorities of the resource-intensive manual contact tracing and made it possible for timely large-scale outbreak containment. The resources saved by leveraging technology can then be used for taking care of vulnerable populations and those without access to a mobile phone.

#### 5. How would a “rapid response” occur, and what would precipitate such a response?- e.g. schools, care homes, other localities?

Hong Kong – On 22^nd^ January, an individual from Wuhan reported respiratory symptoms, and a second suspected case reported the next day. Both cases received medical care, being placed in isolation and later tested positive for Covid-19. Contact tracing started immediately and travel history of patients were retieved and published online. All their close contacts, such as passengers seated close by, taxi drivers, were subject to quarantine at the Lady MacLehose Holiday Village, converted as a quarantine centre to host contacts of confirmed cases. A hotline was set up to answer public enquiries, especially for passengers on the same train/flight.(40)

Singapore – proactive contact tracing and cluster identification are two of features of the country’s Covid-19 response. Health professionals are trained to identify potential clusters through putting forward relevant questions to Covid-19 patients. The Ministry of Health works with hotels for quarantine sites and companies for information such as CCTV footage to track cases.(40)

##### Infectious control

Inadequate infectious control during isolation or quarantine may increase the transmission of SARS-Cov-2. Close quarter isolation was proven ineffective by the Xu et al(41) study of the Diamond Princess. The virus spread to 634 passengers despite the contact tracing(42) and safety measures(43), leading the authors to believe that aerosol contamination was possible in confined complexes. This was contradicted by Wang et al in a hospital study of transmission through air, sewage, surfaces and personal protective equipment in which swabs tested positive only for sewage with virus cultures not being found.(44)

##### Nosocomial infection

and cross infection of doctors, as reported at the Sheffield Teaching Hospitals NHS Foundation Trust, of 1,533 symptomatic healthcare workers, 18% were positive for SARS-CoV-2.(45) It was estimated that a third of staff had completed a shift while symptomatic, emphasising the need for regular and efficient testing for healthcare workers, who have a high risk of infection, for the protection of vulnerable patients and civilians.

To control nosocomial infection, Taiwan implemented nationwide enhanced Traffic Control Bundling (eTCB) in hospitals whereby infection was controlled with a combination of triage prior to hospitalisation, separation between risk zones, strict PPE use and hand disinfection checkpoints.(23, 46) Risk zones were divided into contamination, intermediate and finally, clean. As droplet and fomite transmission have been observed for COVID-19 inside and outside hospitals, containment of nosocomial transmissions with eTCB. This finding was built on research by Yen et al in which SARS infection among HCWs was transmitted to 2HCWs (0.03 cases per bed) in the eTCB hospital compared with 50 probable cases (0.13 cases per bed) in the control group.(47) This strategy was implemented across Taiwan on the 21^st^ May, 2003 and within two weeks the epidemic was under control.

#### 6. How would an app be assimilated in light of the above?

The Singaporean government has developed the app “Trace together”, recording other users who have been in proximity to a smartphone user via Bluetooth. After that user is found to be positive, individuals at risk are contacted directly.(26, 48) The EU PEPP-PT coalition proposed to develop a privacy-friendly contact tracing apps with the use of matching Bluetooth signals, based on the Singaporean “TraceTogether” app.

The Korean Ministry of the Interior and Safety has developed a mobile phone application named “self-quarantine safety protection” App that monitors the location of the quarantined user, informs health authorities, allows the user to report on their symptoms, and health officials can evaluate if a test is needed.(26)

#### 9. What are the barriers to & enablers of being tested, reporting contacts & isolating as a result of being contacted?

##### Logistics

###### Testing

With the United States in mind, Parmet et al. encouraged the promotion of “free testing” in order to reach poorer communities and eradicate the infection.(28) Although the UK already has free testing for adults, few are aware of their availability and necessity in combatting COVID-19. Mark et al. reviewed the feasibility of a mobile community testing team in Scotland and reported a barrier of a relative lack of guidance in infection control for testing in the community, to minimise cross-transmission risk. Other barriers included shortage of staff and long travel times.(49)

###### Contact tracing

Validity and reliability of information recorded by the app may not be accurate or precise due to technological limitations.(48) A high coverage is needed for an app to be effective (a coverage of 60% – 75%), which may be difficult to reach on a voluntary basis, for issues such as mobile storage data, operating system, battery power to support constant Bluetooth activation, young children and senior citizens may not carry or own a personal smart device, and individual willingness. In the UK, only 47% of individuals who are 75 years and older uses internet.(50) It is essential to cater to their needs with the efforts from the outbreak control teams and community volunteers, such as the NHS Volunteer Responders in the UK.(31)

###### Isolation

Logistical challenges increase in providing food, sanitation, transport(51) and care for individuals living in the restricted zone, especially if the epidemic rises to regional or national level.(28)

##### Public awareness and Communication

The number of tests carried out during an emergency will depend on the public perception of the reliability of testing services and the effectiveness of communication of actions that can be taken. It is also necessary to make the availability of kits and location of testing stations accessible to under-privileged members of the public who may not have access to a smart phone or basic supplies. Ethnic minorities should be made aware of the higher death rates statistically observed in members and encouraged to test early. Regular press conferences held by the central outbreak control team, briefing on the progress of the pandemic, changes in policies, and correcting misinformation(23) can increase public awareness of the need of TTI and clarify rumours generated from knowledge gaps and uncertainties.(52)

The internet and social media have a strong impact on isolation behaviour which has grown with increased internet use since the COVID-19 lockdown. Farooq et al. tracked the effects of social media, news websites and emails as well as the living situation on the individual-level intention of self-isolation during the pandemic, using a 225-member survey.(53) It was found that while frequent social media use contributed to information overload and cyberchondria, it increased propensity for self-isolation. McNeill et al similarly studied the effects of tweets and found that social media played a role in the motivation to conform to health measures.(54) To motivate members of the public to self-isolate in a healthy way, a combination of lowering perceived response costs and clear information about the severity of risks should be implemented.(53) It is also suggested that messages of severity of disease be disseminated via reliable official networks such as news, journalism and government outreach while social media focuses on hopeful messages and recommended health measures.

##### Legality

In Singapore, the Infectious Diseases Act provides legal footing to prosecute non-compliance with consequence of fines and/or imprisonment.(24) Similarly, the Communicable Disease Control Act in Taiwan mandates the government to implement effective measures in Covid-19 controls including TTI.(23)

In the US, the Public Health Service Act empowers the Surgeon General (delegated to the Centers for Disease Control and Prevention (CDC)) to enforce quarantine and isolation and provide medical care for detained individuals. Court precedents establishes that quarantines cannot be invidious racially, and governments are required to have a strong basis for imposed restrictions.(28)

##### Public cooperation

The effective control of the highly contagious Covid-19 epidemic replies on the collaboration of individuals to voluntarily provide their information and comply with preventive measures. The potential undesirable outcome in telling the truth, such as adverse immigration consequences for noncitizens, deportation of unlawful immigrants or refuse entry for passengers with a fever, can be a barrier for these individuals, who may be at high risk of infection, to seek care or provide accurate information.(28) Special protection of personal information may be applied with the discretion of the outbreak control team, with a goal to facilitate trust, prioritise care and encourage individuals to cooperate, to ultimately prevent further transmission of the disease.

To ease the economical pressure and uncertainty during large-scale quarantine, governments have established different scheme or acts to support the individuals and families. In the US, the government passed the Families First Coronavirus Response Act, including paid sick leave and unemployment insurance.(55) In the UK, national job retention schemes and financial support schemes have been rolled out early to support individuals and businesses during Covid-19 epidemic.(55)

### A case study on test, contact tracing, testing, and isolation policies for Covid-19 prevention and control in Taiwan

On 31^st^ December 2019, through its communicable diseases news surveillance system, the National Health Commend Center in Taiwan received the information of susceptible SARS-like pneumonia cases in Wuhan. A press conference was hold on the same day and inbound passengers health monitoring started.^1^ In three weeks, on 21^st^ January 2020, the first Covid-19 case in Taiwan was detected through inbound passenger quarantine. The Covid-19 response in Taiwan prioritises on comprehensive border control, effective contact tracing, testing and isolation (TTI), aided by technology and data linkage. The strategy stops the transmission of SARS-Cov-2 without a lock down or mass testing. Starting from 12^th^ April, 2020, there has been no inland Covid-19 case, and since 8^th^ of May the government has gradually eased disease control measures. As of June 8^th^, most restrictions for Covid-19 control such as mass gathering and care home visit are relaxed, whereas the public to practise personal prevention measures of **washing hands, wearing facemask and social distancing**. Between 21^st^ January to 11^th^ June 2020, Taiwan has a total of 443 Covid-19 cases and 7 of them died.

The case study used the Covid-19 response in Taiwan to answer the questions raised by the independent Sage committee, supplementing the literature review. The start and the end of the initial Covid-19 urgent response in Taiwan may inform the UK policy. The implication is facilitated by the similarity of the two countries in their universal health care (single-payment systems, tax-based and full-country coverage) and practice (clinical guidelines, prevention and quality of care initiatives) (Supplementary Table S1).

#### 1. How would a 24 hr turn-around from sampling to result actually work?

##### 1–1. infrastructure and procedure of a sampling-to-results process that is 24 hours or less

The laboratory RT-PCR assay time from receiving the sample to obtaining the result is about 4 hours. The 24 hours turnover thus includes the delivery time of the sample to the certified laboratories. In January, there were 15 laboratories certified for Covid-19 RT-PCR test, covering different administrative areas in Taiwan. Each laboratory has specified time window to receive samples and release results twice daily. **Change in Covid-19 case definition** the case definition of Covid-19 changed frequently as the epidemic evolved (figure 1), and each time the change introduced a surge in cases reported and testing needed for suspected cases. As part of the contingency plan for large-scale outbreak, the government continues to scale-up testing capacity by facilitating regional hospitals to establish a Biosafety level-2 negative pressure laboratory (BSL-2) and strengthening the local testing network. There has been a gradual increase in the number of certified laboratories, (Figure 2) and as of 2^nd^ of June, there was 45 laboratories, within which 11 are designated Biosafety level-3 laboratory and 19 of them can perform tests at any time, bringing a daily testing capacity to around 6,000.

As no new Covid-19 cases reported in Taiwan since April, the mandated testing for suspected cases or individuals at high risk of infection accounted for about 4% of the total testing capacity. Therefore, starting from May, the government has expanded the testing scheme, and the public can pay to have their samples taken at 162 designated community collection and inspection centres (the CDC Taiwan testing centre map for the public https://antiflu.cdc.gov.tw/ExaminationCounter).

##### 1–2. testing during emergency

In addition to the testing network, the Industrial Technology Research Institute developed a portable RT-PCR kit (size of a soda can) with time from sample to result about 60 minutes.^2^ For the large-scale outbreak when it requires mass testing with rapid turnaround, the point of care testing (POCT) system can be implemented. The portable equipment enables testing on site and can shorten the turnaround time to 1–2 hours. Using synthetic and monoclonal antibodies for rapid testing is being developed.

##### 1–3. minimising false-positive and false-negative results

The CDC in Taiwan designs and provides the primer for SARS-Cov-2 RT-PCR testing, according to the guideline of WHO or US CDC.^3^ The assay is highly sensitivity (E-gene results: 3.7–9.6 RNA copies/rxn).^4^ However, as at the initial stage of the infection, the viral shedding can be too low to be sampled, resulting in a false negative result due to sampling. Thus for susceptible cases, a **Two-sample diagnosis** rule is applied; after the first sample (nasal or throat swab), the second sample is taken after 24 hours to ensure accuracy.

#### 2. What local governance and partnership structures are required, and what would a local “outbreak team” look like?

The **community healthcare network** began to deliver the community Covid-19 prevention in Taiwan from January 26^th^, 2020, 5 days after the first confirmed case reported.

The building blocks of the community healthcare network are **Public health/health service centres** in each township. Their roles are to monitor, investigate, and report Covid-19 cases, to follow up on and provide care for those in home isolation or quarantine, and to deliver disease prevention supplies for the community.

The work of public health/health service centres is supervised and supported by the **local government**. In the Covid-19 response, each local government establishes a care service center, integrating local police, borough office, health centres/hospitals in its administrative area, to coordinate information and resources to follow up cases and provide individuals in home quarantine and home isolation with services including daily follow-up, psychological counselling, medical care assistance, meal delivery, transportation, garbage collection, and hotline services. For individuals breaking home isolation or quarantine regulations, the local authority is also in charge of finding, advising and issuing fines. The steps the local government care service center takes for managing Covid-19 is summarised in figure 3. Local health authorities and specialised public health physicians carry out outbreak investigation.

The roles and coordination of the local outbreak team members are specified in the articles 5 and 6 of the Communicable Disease Control Act.^5^ Table 1 summarises the members of the case management team, their roles, procedures and how they work together in carrying out home isolation, home quarantine, and self-health management. It is notable that individuals undergoing home isolation or quarantine are required for an additional 7-day self-health management at the end of their two-week isolation or quarantine period.

**Figure 1:**
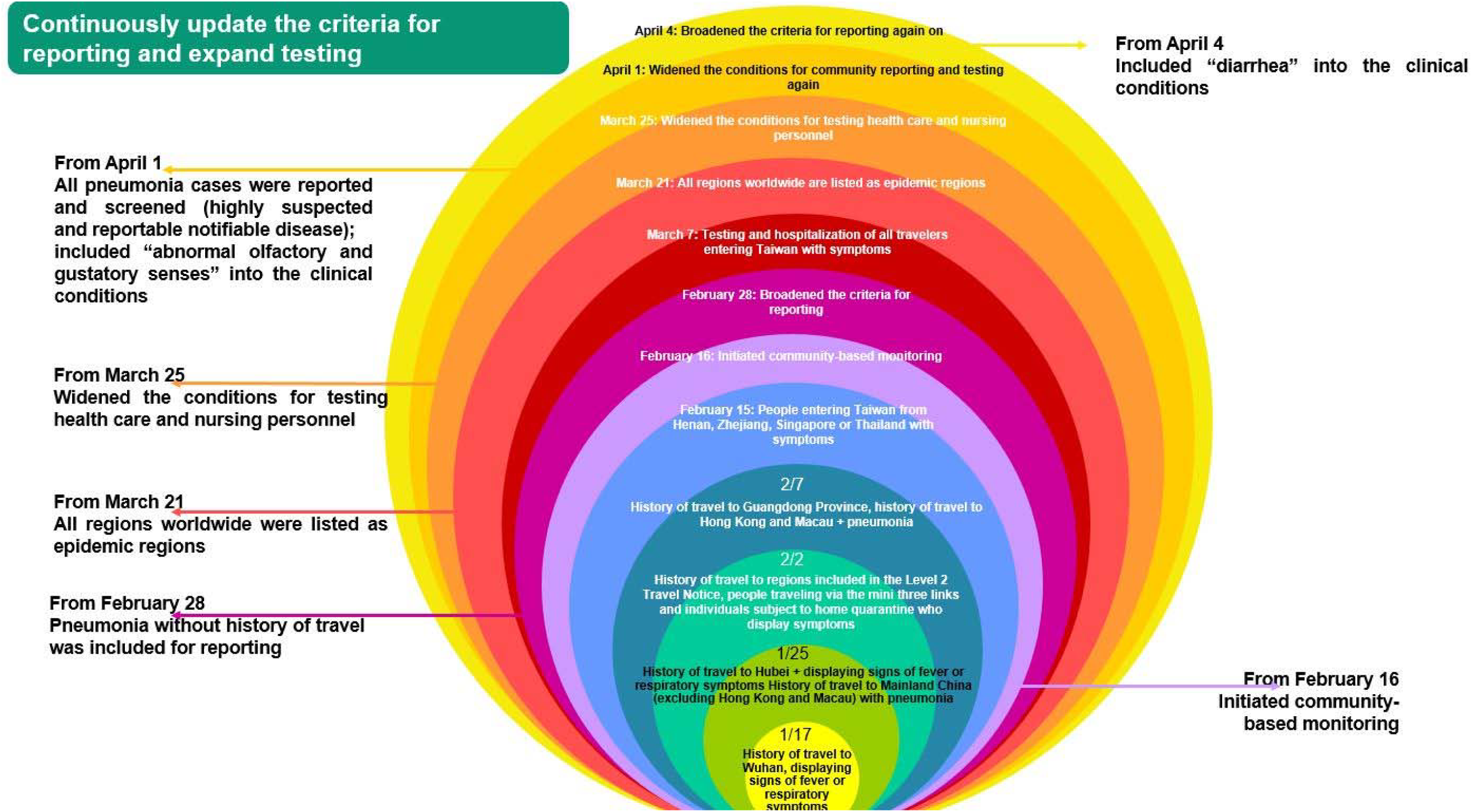
The evolution of Covid-19 reporting criteria and expansion of testing. (adopted from the display from the webpage: https://covid19.mohw.gov.tw/en/cp-4775-53739-206.html.

**Figure 2:**
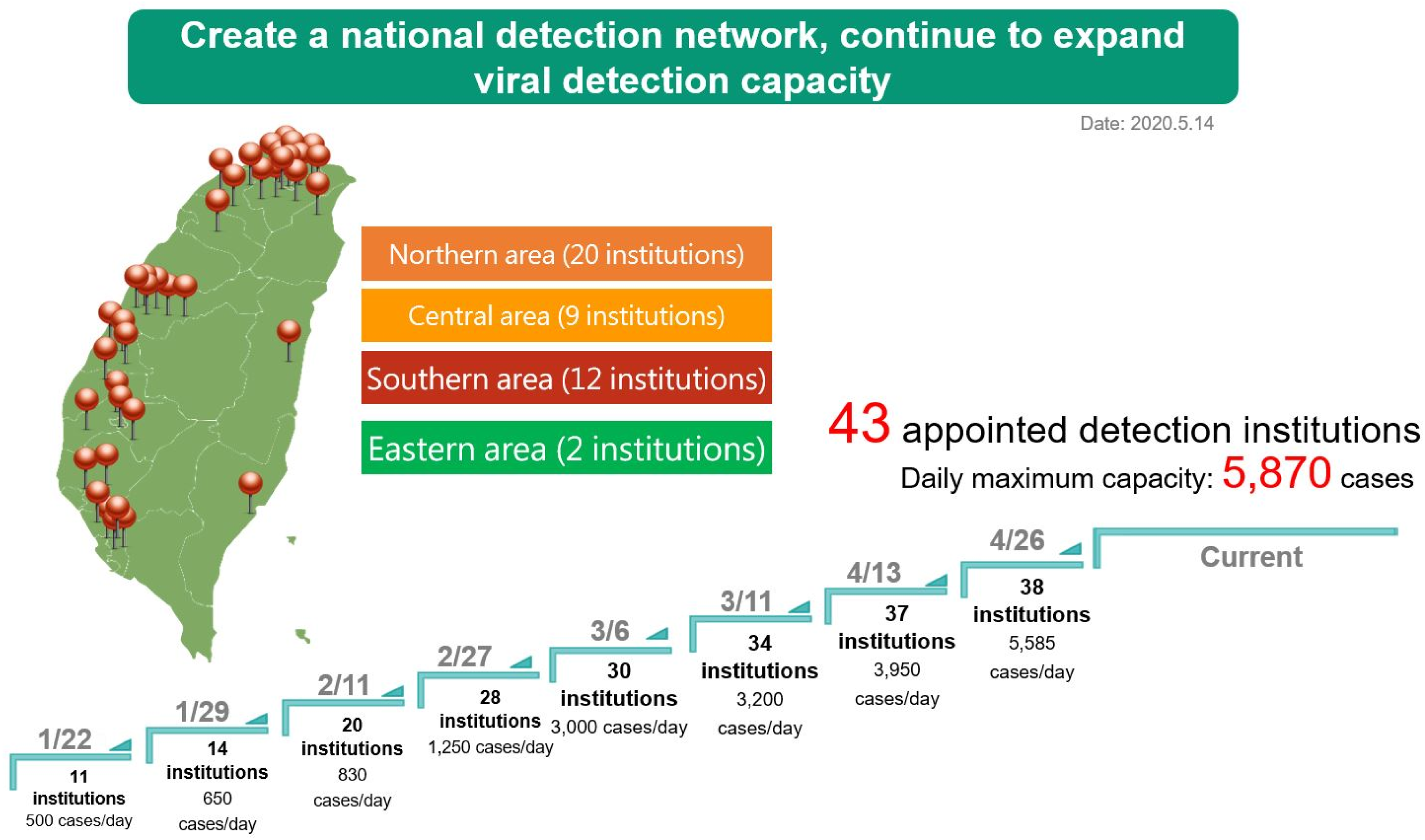
The increment of Covid-19 testing capacity in Taiwan (adopted from the display from the webpage: https://covid19.mohw.gov.tw/en/cp-4788-53906-206.html).

**Figure 3.**
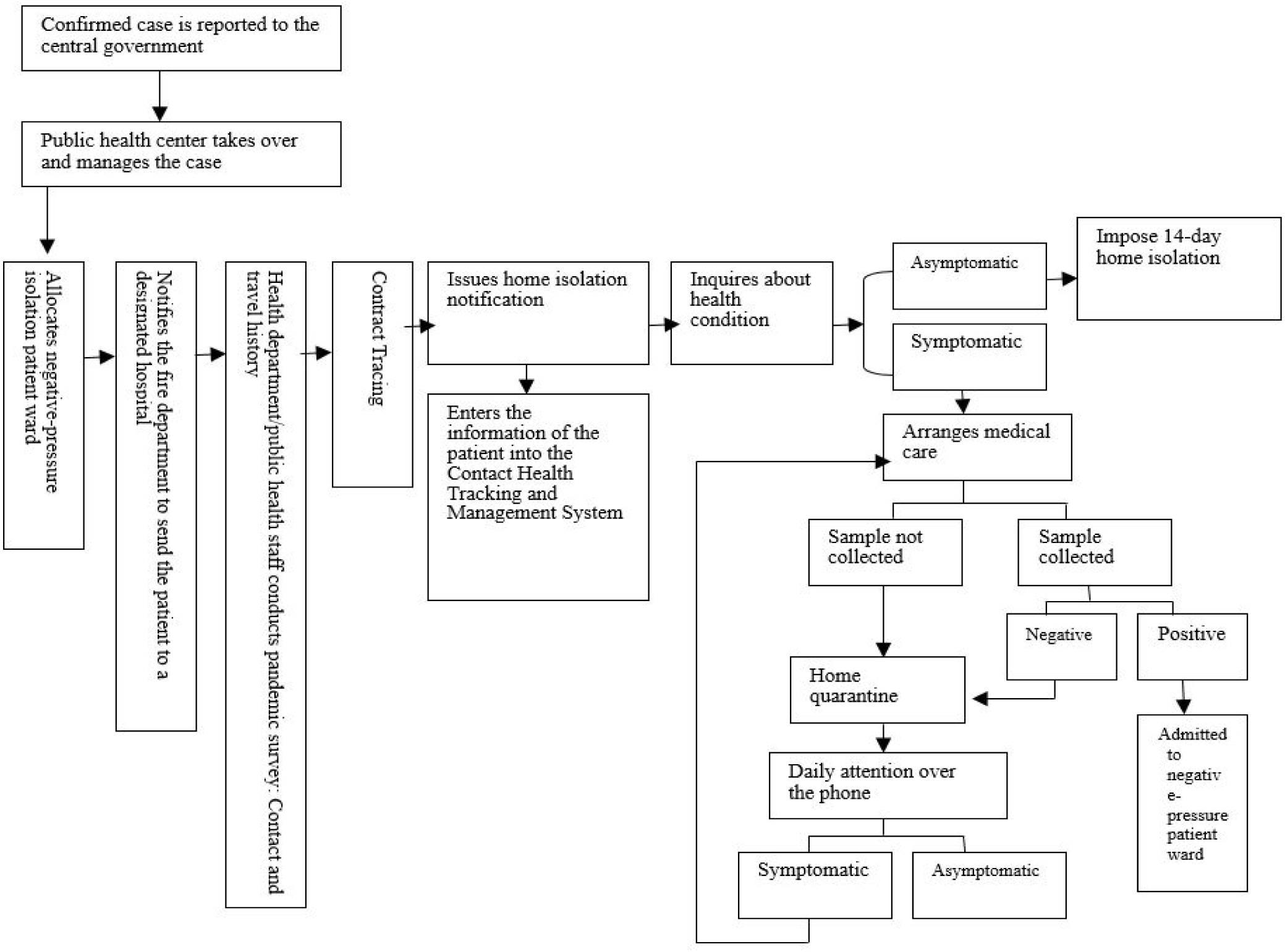
Procedures for Covid-19 management. (adopted from the display from the webpage: https://covid19.mohw.gov.tw/en/cp-4781-53799-206.html.

**Table 1:** the role, procedures and collaboration of local outbreak control team for home isolation, quarantine and self-health management in Taiwan.

| Intervention | Home Isolation | Home Quarantine | Self-health management |
| --- | --- | --- | --- |
| Groups of persons | Persons who had contact with confirmed cases | People with travel history | 1. Reported cases who have tested negative and met criteria for being released from isolation<br>2. People reported and tested for COVID-19 under "COVID-19 Community-based Surveillance" |
| Responsible authorities | Local health authorities | Local civil affairs bureau or borough chief | Central/Local health authorities |
| Enforcement | Home isolation for 14 days<br>Active monitoring twice a day | Home quarantine for 14 days<br>Active monitoring once or twice a day | Self-health management for 14 days |
| Notes concerning respective measures | Health authority will issue a "Home (Self) Isolation Notice"<br>Health authority shall check health status of the individual twice a day<br>During the home isolation period, the individual is to stay at home (or designated location) and not go out, and may not leave the country or use public transportation<br>Symptomatic individuals will be sent to the hospital for medical attention<br>Individuals not adhering to the CECC's prevention measures will be penalized under the Communicable Disease Control Act and be forcibly placed<br>After the home isolation period ends, the individual should conduct an additional 7-day period of self-health management | Where the relevant authority has issued a Novel Coronavirus Health Declaration and Home Quarantine Notice, the individual is to wear a surgical mask and return home for home quarantine<br>The local borough chief or borough clerk shall call the individual every day during the 14-day period to ask about the individual's health status, and shall record the information obtained<br>During the quarantine period, the individual is to stay at home (or designated location) and not go out, and may not leave the country or use public transportation<br>Symptomatic individuals will be sent to designated medical facilities for tests; the relevant health authority will also begin active monitoring<br>Individuals not adhering to the CECC's prevention measures will be penalized under the Communicable Disease Control Act and be forcibly placed<br>After the home quarantine period ends, the individual should conduct an additional 7-day period of self-health management | Asymptomatic individuals are to avoid public places, postpone any non-urgent medical care or examination, always wear a medical mask when going out, wash hands frequently, follow respiratory hygiene and cough etiquette, and take temperature twice a day, once in the morning and once in the evening.<br>Individuals with fever or respiratory symptoms such as coughing or running nose are to wear a medical mask, seek medical attention immediately and not to use public transport; inform the physician of your contact history, travel history, and whether anyone else has similar symptoms; wear a surgical mask while returning home and avoid going out; and keep 1 meter away from others when talking to them.<br>After being tested for COVID-19 and returning home, individuals are to stay at home and not to go out before receiving results<br>Medical personnel are to halt work and not to come to work temporarily |
| Legal basis | § Article 48, Communicable Disease Control Act<br>§ Paragraph 1, Article 15, Special Act for Prevention, Relief and Revitalization Measures for Severe Pneumonia with Novel Pathogens | § Article 58, Communicable Disease Control Act<br>§ Paragraph 2, Article 15, Special Act for Prevention, Relief and Revitalization Measures for Severe Pneumonia with Novel Pathogens | § Article 48, Communicable Disease Control Act; Article 58, Communicable Disease Control Act<br>§ Article 67, Communicable Disease Control Act; Article 69, Communicable Disease Control Act |

Central Epidemic Command Center
www.cdc.gov.tw
Communicable Disease Reporting and Consultation Hotline : 1922

In response to the COVID-19 crisis, the central government established the **Central Epidemics Command Center**^6^ (CECC) on January 20^th^ as well as the **expert advisory committee** providing scientific and clinical advice directly to CECC. The role of the CECC is to coordinate different ministries for a joint Covid-19 response, set national regulations for disease prevention and control, supervise and support local outbreak response

#### 7. Adherence to isolation and local support needs

##### 7–1 the ways to know if a suspected case is adhering to isolation

Within the first few hours of the starting of self-isolation or quarantine period, staff from local health authority or borough will phone the suspected case(s) to communicate necessary steps. Subsequently, the staff rings the individual twice daily (home isolation) or once per day (home quarantine) to follow-up. If the call is unanswered, a home visit will be arranged to check if the individual needs further medical care.

###### Electronic fencing

“Electronic Fencing” uses base station triangulation technology to approximate the holder of the mobile phone and automatically issues a reminder when it is beyond the designated isolation or quarantine area. When a mobile phone is switched on, the SIM card will connect with the base station, and the device actively searches for several base stations in the vicinity to connect with the closest one with the strongest signal. The signal strength of different base stations will change according to the location of the mobile phone. Thus, by selecting three base stations, the location of the phone can be triangulated and approximated.

The mobile phones retained by individuals undergoing home isolation or quarantine are registered within the range. Once they leave the range, it will automatically issue an alert to the holder (much like receiving roaming text messages automatically when going abroad), as well as the local government, health authority and local police. Based on the information, the responsible staff can visit the roaming case.

The electronic fencing effectively improves the adherence to home isolation or quarantine, as the probability of going beyond the range dropped from about 30% to 0.3% from its initial launch to the present.

##### 7–2 support for individuals in self-isolation

1. As described in section 2, the local government care service center provides a spectrum of services to support individuals during home-isolation.
2. At the beginning of the 14 days, local government prepares and delivers to the individual under self-isolation a package with food, facemasks, nutritional supplements, and entertainments (book, prepaid movie account, plants).
3. There is a daily phone call from the local health authority and borough to make sure if the individual undergoing home quarantine or isolation is doing okay. They speak to practically all suspect cases, including small children who can talk.
4. To support the individual under self-isolation and to ease the workload of the community outbreak team, the government collaborates with telecommunication companies (HTC DeepQ, Line) to develop a chatbot for individuals serving home isolation or quarantine. They can report their daily health conditions to health authority via texts and receive advice, such as on the 13^th^ and 14^th^ day of home isolation or quarantine period, they will receive a reminder on the 7-day self-health management after the completion of isolation period.
5. There is an allocation of 1,000 NTD per day per diem during home isolation or quarantine.^7^

#### 4. How would real-time data management, linkage of datasets, and dashboards be developed, and who would “own this”?

##### A. Epidemiology Surveillance System

According to law,^8^ health authorities are mandated establish an epidemiological surveillance and advance-alert system for communicable diseases. The components include:

1. National Notifiable surveillance and advance-alert system;
2. Laboratory surveillance and advance-alert system;
3. Sentinel medical institution surveillance and advance-alert system;
4. School-based surveillance and advance-alert system;
5. Nosocomial infection surveillance and advance-alert system;
6. General public surveillance and advance-alert system;
7. Disease control material surveillance and advance-alert system;
8. Populous institution surveillance and advance-alert system;
9. Symptom surveillance and advance-alert system;
10. Real-time outbreak disease surveillance and advance-alert system;
11. Other surveillance and advance-alert system of communicable diseases.

The information of communicable disease identified by these systems is used for real-time outbreak risk assessment of the health authorities.

##### B. Digital Covid-19 prevention system

The digital Covid-19 prevention system in Taiwan includes a QR-code-based health data collection in the airport for inbound passengers. The data are linked to the “epidemic prevention tracking system” used by the health authorities and local police to inform them of the incoming cases for them to record follow-up information. For travellers arriving at Taiwan without a Taiwanese phone number, a mobile phone will be provided to them for contact during home isolation or quarantine. During home isolation and quarantine, the “electronic Fencing System”, described earlier, facilitates adherence. (Figure 4)

Such information is lawfully collected by the health authorities and government for outbreak control.^9^However, the authorities can only retain the information for 28 days (the electronic fencing system retains the information for only one day) and required by the CECC regulation to subsequently eliminate and destroy the data for the protection of individual data privacy, confidentiality and security.^10^

The digital Covid-19 prevention system is linked to National Health Insurance MediCloud System, a cloud-based medical information sharing platform for medical professionals to gain updated clinical data of the patient seeking care, including testing, imaging, medication, and treatment results.^11^ Linking the Covid-19 TTI information to MediCloud provides medical staff with real-time information on the patient’s Covid-19 travel and contact history, home isolation and quarantine status to provide care following appropriate infectious control measures, for the protection of health workers and other patients.^12^ Real-time data sharing and linkage is conducted via a closed exclusive network system (virtual private network (VPN) for data security.^13^

##### C. Example of digital Covid-19 prevention system leverage big data and health data linkage – Diamond Princess cruise ship contact tracing.^14^ (Supplementary session two)

The index case on Diamond Princess Cruise ship was reported on 20^th^ January 2020. On 31^st^ January, there were 3000 passengers from the Diamond Princess cruise disembarked at Keelung harbour in Taiwan for a 1- day tour. Soon afterwards, on 5^th^ February, the cruise ship reported a Covid-19 outbreak.

To manage the potential risk or large scale Covid-19 outbreak, the Taiwanese government used geopositioning analysis on mobile sensor data, cross validated with other surveillance data, to identify 627,386 potential contacts of the 3,000 Diamond Princess passengers. The telecommunication companies sent text messages to these potential contacts with the information on home quarantine, self health management and subsequent RT-PCR testing if developing symptoms. The contact data were linked to a National Health Insurance claims dataset, for the health authorities to follow their health outcomes. Three weeks afterwards, as of 29^th^ February, no Covid-19 incident was reported from the contacts.

#### 5. How would a “rapid response” occur, and what would precipitate such a response?- e.g. schools, care homes, other localities?

The overarching strategy to prevent Covid-19 community transmission is published by the CDC Taiwan on 6^th^ April, 2020, listing the following 12 strategies:^15^

A. At the individual and family level 1) health promotion and risk communication, 2) isolating Covid-19 cases, 3) isolating close contacts (with methods such as home isolation/quarantine, institution quarantine and workplace quarantine).
B. At the community level 4) regional quarantine, 5) enhancing infectious disease control or suspending public gathering, 6) enhancing infectious disease control in public transportation, 7) school suspension or closure, 8) enhancing infectious disease control in public areas and/or retailing stores, 9) rapid containment, 10) sheltering, 11) domestic travel restriction, 12) cordon sanitaire.

##### 5–1. Hospitals

The contingency plan to prevent coronavirus infection in hospitals has been established according to the principles of Chemical, biological, radiological and nuclear (CBRN) defence and protective measures,^16^ after the nosocomial infections during the SARS outbreak in 2003. It includes the following three main principles.

1. the division of hospital areas according to Covid-19 infection risk as contaminated (such as negative pressure ward, ICU, CCU), intermediate and clean zones;
2. triage of patients (patients with Covid-19 like symptoms will be managed and treated in designated areas separated from other non-Covid-19 medical care activities);
3. medical staff divide into set teams and locations to deliver care, and change in teams and care station is set to minimum. (As in a submarine, the compartments can be completely separated when seawater enters.)

The infectious control aimed to minimise the fomite and aerosol transmission. During SARS, hospitals implemented the contingency measures reduced the risk of SARS nosocomial infection by half as compared to hospitals without the preventive measures in place.^17^

###### Hospital covid-19 cluster infection contingency protocols

If two Covid-19 clustering occurred within 14 days in a hospital, followed by a third cluster infection, the hospital to report to corresponding health authority to conduct outbreak investigation. Judging from the findings of the investigation, the hospital and health authority can initiate the “operational control” or “clearing control” protocols for 28 days.^18^ Operational control minimize all not-essential care activities to apply thorough disinfection, whereas cleaning control moves all patients and staff from the contaminated area to an allocated area or designated hospital within the medical network^19^ to continue care. Hospitals to prepare for contingency with simulation practices.

On the other hand, due to the severity of Covid-19 pandemic worldwide, from 23^rd^ February, restriction on international travel has been placed on medical staff and social care workers, with reimbursement. In hindsight, the decision protected and maintained the clinical care force during the peak time of the epidemic (mid-March in Taiwan, due to the retuning citizens infected with Covid-19 from abroad).

##### 5–2. Care home

The infectious control^20^ and contingency plan^21^ for long-term care homes are similar to the ones for hospitals. A restriction on visitors have been placed on 17^th^ April but relaxed two weeks afterward by 1^st^ May, visitors to follow regulations on registration, reduced frequency and personal prevention measures of hand-washing, wearing facemask, and social distancing (1.5 meters indoors and 1 meter outdoors).

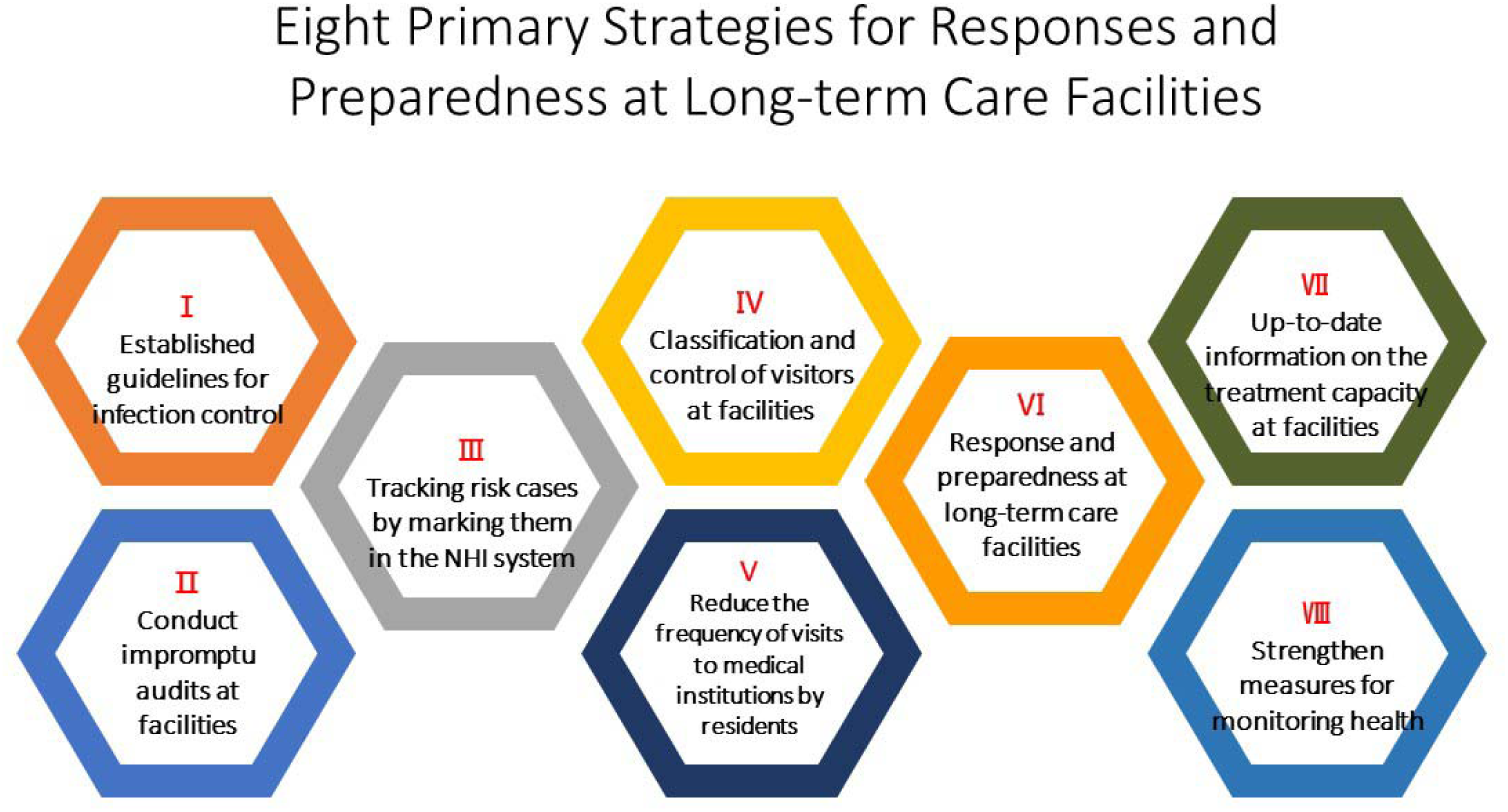
(adopting from https://covid19.mohw.gov.tw/en/cp-4787-53905-206.html)

##### 5–3. School

The ministry of education together with CDC Taiwan published the criteria for school suspension in response to Covid-19.^22^ The duration of suspension is 14 days.

1. For schools other than universities:

1. When one teacher or student identified as a confirmed case by the Central Epidemic Command Center (CECC), the class is suspended for 14 days.
2. When more than two teachers and students in a school are identified as confirmed diseases by CECC, the school is closed for 14 days.
3. When one-third of schools in a borough are closed due to Covid-19, all schools in the borough are closed for 14 days.
4. To prevent cluster transmission, from 16^th^ March until the end of the Spring semester in 2020, students and teachers up to high schools have been subject to restriction on international travel, with special considerations given to exceptional cases.
2. For universities:

1. when a teacher or student identified as a confirmed case by CECC, all courses taught by the teacher or taken by the student are suspended for 14 days.
2. When more than 2 teachers and/or students are confirmed cases by the CECC, the university is closed for 14 days.

#### 6. How would an app be assimilated in light of the above?

As a part of the contingency plan, the government collaborates with telecommunication companies to develop a health report App, integrating the digital Covid-19 prevention system in Taiwan (as described in the answer to question 4 and question 7 above).

Another App developed is a social distancing App, using bluetooth device signal to estimate the physical social interactions out of devices close to individual. The App meets GDPR regulations and generates anonymous hashed ID history stored at the device for up to 28 days.^23^

#### 9. What are the barriers to & enablers of being tested, reporting contacts & isolating as a result of being contacted?

##### 9–1. enablers

1. The effective control of Covid-19 indicated by low incidence and mortality is an ongoing encouragement for the country, which has gone through SARS, to follow the regulations.
2. The care and support provided by the local outbreak control team, individuals undergoing home isolation or quarantine have expressed their appreciation of being contacted once or twice daily by the health authorities and borough staff. The warm gesture of 14-day stay-at-home package prepared and delivered by the local government to individuals undergoing isolation or quarantine also helps.
3. The public trust developed with the leadership of CECC, central and local teams, with the transparency of information, effective communicating of risk, and the care and humanity shared by the CECC and teams working on Covid-19 prevention and control. In addition, the hard work, care and kindness given by the medical staff. The public consider that if they follow the guidance, they will be safe, and if they are unfortunately getting sick, they will be well looked after.
4. The precision, efficiency and safety of the Covid-19 prevention system. At the airport, digital “entry quarantine system” has shortened the time from entry to care from 19 hours to 4.5 hours. The electronic fencing has greatly reduced the proportion of individuals going out of home isolation or quarantine from 30% in January to 0.3% currently and may significantly reduce potential community transmission.
5. The per diem of 1,000 NTD each day for home isolation and quarantine and the fine of 100,000 –1 million NTD for violating the home isolation or quarantine regulations.^24^

##### 9–2 barriers

Perceived shortage of personal prevention materials and false information can disturb the public with uncertainty and worry.

Taiwan is a diverse society with major immigrant populations. To overcome the potential language and culture barriers. The reporting and consultation hotlines are available in four commonly speaking languages and with telecommunications device for the deaf. All health education materials and important forms, such as notice for home isolation or quarantine, are translated into multiple languages, and news communicated in different dialects.

###### The coping strategy

Overcoming the shortage of personal prevention materials – make your own.

###### Masks

In early February, Taiwan’s daily mask production capacity was only 2.71 million pieces. By mid-May, 114 production lines were added with a daily output of 19 million pieces. The main contributors are the mask national team (production), the military (production), Taiwan Textile Research Institute (coordination), the post office (distribution) and eMask platform (fair distribution – purchase with the health insurance 1C card).^25^ A total of 700 million masks were distributed with low price to the public since February, and currently there are 350 million of inventory of medical and surgical masks. 220 million masks were distributed free to medical professional and staff.

###### PPE

Before Covid-19, Taiwan imported all PPEs from abroad. To address the need of PPE, the government collaborates with the textile industry and material factories to produce PPE in house. The first production of 1 million PPE is about to complete.

###### Alcohol disinfectant

The government recruited the Tobacco and Liquor Corporation and Sugar Corporation to produce alcohol disinfectant since February, and 19.3 million bottles have been delivered for medical or household use since.

###### False information control

According to law,^26^ individuals who disseminate rumours or false information regarding the Covid-19 epidemics, causing damage to the public or others, can be sentenced to imprisonment and/or a fine up to NT$3 million.

#### 3. What local commissioning arrangements are needed to facilitate this?

Local commissioning may be specific to the UK context. In Taiwan, the collaboration between the central and local government and hospitals to establish the communicable disease medical network may hopefully be helpful (Regulations Governing Operation of the Communicable Disease https://law.moj.gov.tw/ENG/LawClass/LawAll.aspx?pcode=L0050014 The budget document can be included upon request.)

#### 8. Messaging to the public and behavioural incentives^27^

1. Staring from 22^nd^ January, the CECC recruited all television and news channels in Taiwan to broadcast Covid-19 news at least twice every hour to keep the public informed.
2. From 22^nd^ January to 7^th^ June, the CECC team held daily press conference and published at least one daily press release to communicate with the public the progression and control of Covid-19. After the urgent response ended on 7^th^ June, the frequency of CECC press conference is reduced to every Wednesday.
3. The capacity of the Communicable Disease Reporting and Consultation hotline has been increased from the initial 16 staff and 90 lines to 400 staff serving 270 lines, to ensure the public receive a timely response to their inquiries.
4. Information on outbreak, government response and health education materials (in different languages) are provided on various advertising channels, including posters, leaflets, media and social media to communicate truthful information and promote public trust, cooperation and practice on protective measures.

## Data Availability

All data relevant to the study are included in the article or uploaded as supplementary information.

## Supplementary session one: list of questions to synthesise information from the eligible studies in the review

**Figure 1:**
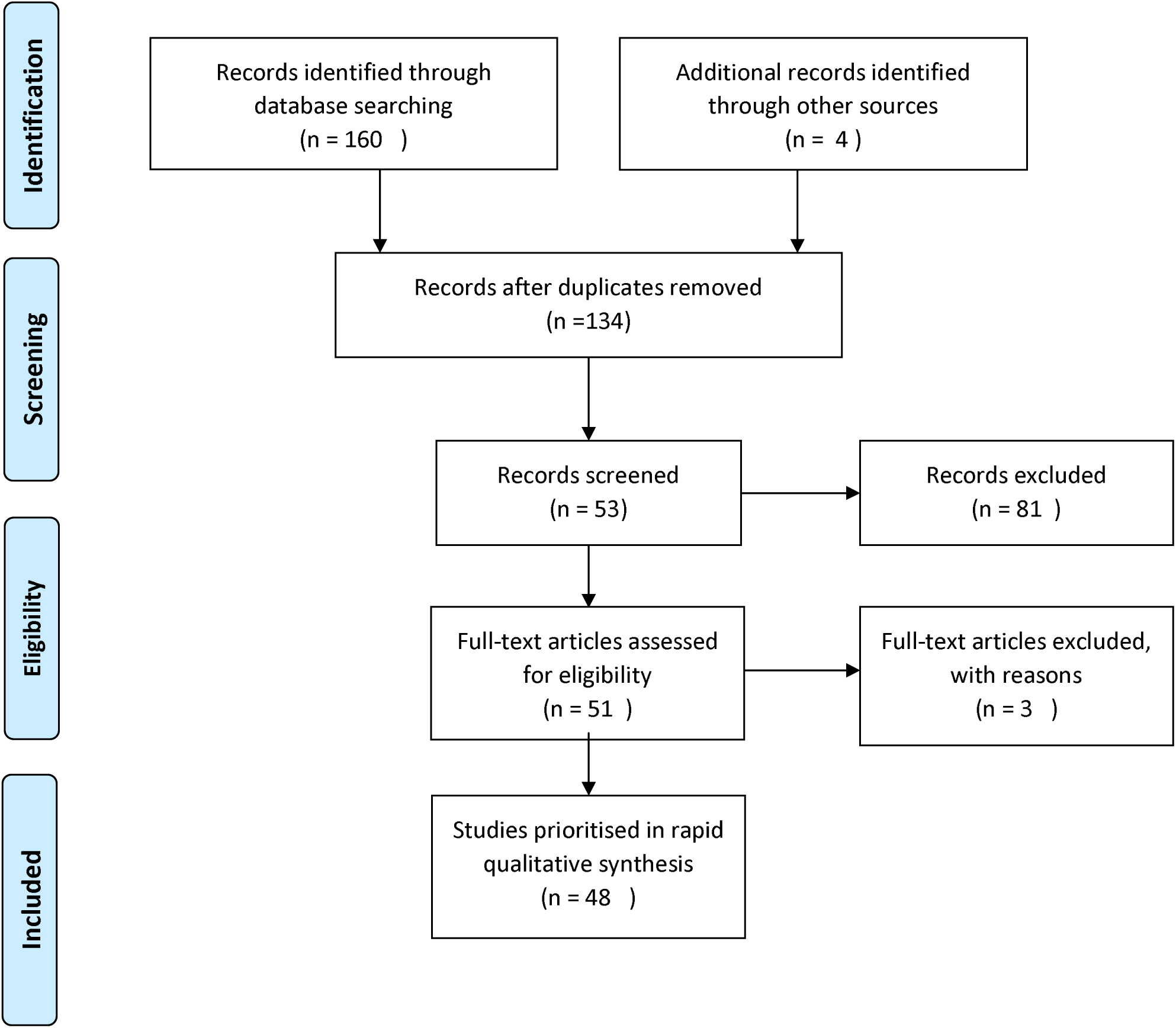
systematic review study flow diagram.

### Study summary

Title

Author

Journal

Research question or objective

Study design

Study population

Sample size

Analysis or Method

Main findings

Implications

Main limitation(s)

### Policy questions

1. How would a 24 hr turn-around from sampling to result actually work? (the report only goes as far as suggesting a 3 day turnaround)
2. What local governance and partnership structures are required, and what would a local “outbreak team” look like?
7. Adherence to isolation, and local support needs
3. What local commissioning arrangements are needed to facilitate this?
4. How would real-time data management, linkage of datasets, and dashboards be developed, and who would “own this”
5. How would a “rapid response” occur, and what would precipitate such a response?- eg schools, care homes, other localities?
6. How would an app be assimilated in light of the above?
9. What are the barriers to & enablers of being tested, reporting contacts & isolating as a result of being contacted?

**Supplementary Table S1.** Population health profile and health care comparison between Taiwan and the UK.

| Characteristics | Taiwan | UK |
| --- | --- | --- |
| Demographics |  |  |
| Population, (million) (2015) | 23.5 | 65.1 |
| Female, % (2011) | 49.9 | 50.9 |
| Population density, persons per km<sup>2</sup> (2013) | 646 | 413 (England) |
| Gross national income per capita, (PPP international \$) (2016) | 48,095 | 42,481 |
| Gini coefficient (2013) | 0.336 | 0.358 |
| Health system indicators |  |  |
| Gross national income spent on health, % (2013) | 6.0 | 9.3 |
| Curative (acute) hospital beds, per 1 000 population (2014) | 3.4 | 2.3 |
| Negative pressure quarantine beds, whole country | 963 | N/A |
| Population Health Profile |  |  |
| Life expectancy at birth, years (2015) | 80.2 | 81.2 |
| Tobacco use among aged 15+ (2013) | 18 | 19 |
| Liters per capita alcohol consumption (2011) | 2.6 (18+) | 9.9 (15+) |

Data source: National statistics, the Republic of China (Taiwan) https://eng.stat.gov.tw/mp.asp?mp=5; National health insurance administration, ministry of health and welfare https://www.nhi.gov.tw/English/; Health promotion administration, ministry of health and welfare Office of National Statistics https://www.hpa.gov.tw/EngPages/Index.aspx, UK https://www.ons.gov.uk; OECD.Stats http://stats.oecd.org/ <sup>1</sup>CECC press conference 2, June, 2020.

## Supplementary session two: mobile geopositioning method with health data linkage for Diamond Princess cruise ship contact tracing

Background of the event is described in the case study 4-C.

### Identify trails of the cruise passengers

As it was impossible to conduct retrospective individual interviews for each passenger, the method used to find out the location of the passengers have visited and the contact was primarily mobile geopositioning, verified with data from vehicle GPS, credit card transaction log, and closed-circuit television (CCTV).

The mobile position data from more than 3000 passengers on January 31, 2020, were obtained from five telecommunication companies in Taiwan. The cruise was docked at the harbour from 6 AM to 6 PM. The contact locations were ascertained with roaming signals with time of exposure over 30 minutes from multiple mobile base stations between 5 AM and 8 PM. The mobile signals registered to the base stations of telecom companies were compared with the data before, during and after the docking of the cruise to retrieve mobile phone numbers of passengers.

The team was then able to depict rough locations marked by the signals of these phones, and about 34% of passengers took shuttle buses for local tours, 5.2% took taxies, the others biked or walked around at the harbour or the nearby area. The travel agency provided the itinerary of the day trip, and with the assistance of the local government, more than 24 buses and 50 taxies had been interviewed to recover the journeys of the 3000 passengers.

### Identifying the Possible Contacts

The team resorted to the mobile position information of passengers to identify the sensors of mobiles from the possible contact persons. Citizens who carried their mobile phone and stayed within 500 meters of the marked locations over 5 minutes on 31^st^ January 2020 were classified as people who possibly contacted the passengers of the Diamond Princess cruise ship. The number of potential contacts was 627,386.

On 7^th^ February 2020, 2 days after the news of cruise outbreak reported, the CECC sent an alert notice using text message through the Public Warning System to remind the 627,386 contacts and activate the mitigation plan. To stop further transmission, the potential contacts were advised to quarantine at home, monitor COVID-19 symptoms (fever, cough, and shortness of breath), and contact health authority to seek medical care if presenting symptoms.

On 9^th^ February, the CECC notified all health care providers of the incident and provided guidelines for management of symptomatic contacts. Health care professionals were advised to test contacts presenting symptoms and provide care accordingly. Health care professionals were also advised to proactively contact health authorities to initiate follow-up of symptomatic contacts.

### COVID-19 Surveillance for Contact Population Using National Health Insurance Claims Data

In order to identify those in the contact population who sought medical care but did not report to public health authorities, National Health Insurance Claims data were used to track the health status of all potential contacts. Those who were hospitalized due to pneumonia were identified. For those who remained hospitalized but had not been tested for SARS-CoV-2, the hospitals were informed of the potential Covid-19 exposure of the patient and screening for SARS-CoV-2 was suggested.

The data was lawfully accessed and linked under the Taiwan Infectious Disease Control Act, for the purpose of containing disease outbreak, authorization or consent to the retrieval of individual information by the relevant government authorities can be waived. On the other hand, the data is to be eliminated after 28 days for personal data protection.

## Footnotes

1 Crucial policy for combating covid-19. Ministry of Health and Welfare. Taiwan, https://covid19.mohw.gov.tw/en/mp-206.html (accessed on June 10^th^, 2020).

2 Breakthrough in COVID-19 Testing: ITRI Develops Nucleic Acid Detection System for 1-Hour Rapid Tests. Industrial Technology Research Institute. https://www.itri.org.tw/english/ListStyle.aspx?DisplayStyle=01content&SitelD=l&MmmlD=617731531241750114&MGID=1071472323705464234 (accessed at June 10^th^ 2020)

3 WHO.Diagnostic detection of 2019 nCoV by real time RT PCR. https://www.who.int/docs/defaultsource/coronaviruse/protocolv21.pdf?sfvrsn=a9ef618c_2USCDC.2019-Novel Coronavirus (2019-nCoV) Real-time rRT-PCR Panel Primers and Probes https://www.cdc.gov/coronavirus/2019ncov/lab/rtperpanelprimerprobes.html

4 Corman VM, et al. Detection of 2019 novel coronavirus (2019-nCoV) by real-time RT-PCR. Euro Surveill. 2020.

5 Communicable Disease Control Act https://law.moj.gov.tw/ENG/LawClass/LawAll.aspx?pcode=L0050001

6 Enforcement Regulations Governing the Central Epidemics Command Center https://law.moj.gov.tw/ENG/LawClass/LawAll.aspx?pcode=L0050025

7 according to article 4 of the Regulations Governing Disease Prevention Compensation During Severe Pneumonia with Novel Pathogens Isolation and Quarantine Periods. https://law.moj.gov.tw/ENG/LawClass/LawAll.aspx?pcode=L0050040

8 Article 3 of the Regulations Governing the Implementation of the Epidemiological Surveillance and Advance-Alert System for Communicable Diseases https://law.moj.gov.tw/ENG/LawClass/LawAll.aspx?pcode=L0050006

9 Personal Data Protection Act https://law.moj.gov.tw/ENG/LawClass/LawAll.aspx?pcode=IQ050021

10 “COVID-19 (Wuhan Pneumonia)” Disease prevention New Life Movement: Guidelines for the Implementation of actual communication.” https://www.cdc.gov.tw/File/Get/t-Xs5DDee2qzBFC1fRXJA

11 Enhanced Patient Safety and Information Security Both through the NHI MediCloud System. National Health Insurance Administration. https://www.nhi.gov.tw/english/News_Content.aspx?n=996D1B4B5DC48343&sms=F0EAFEB716DE7FFA&s=148EAC979AAE3F75

12 Information on departures/transits to high-risk areas is now included in the travel history notification list, thus effectively closing gaps in disease prevention. National Health Insurance Administration. https://www.nhi.gov.tw/English/News_Content.aspx?n=996D1B4B5DC48343&sms=F0EAFEB716DE7FFA&s=00AF96BA7D327FF5

13 Taiwan Can Help – National Health Insurance’s Contribution in Combating COVID-19. Ministry of Health and welfare. https://covid19.mohw.gov.tw/en/cp-4778-53691-206.html

14 Chen CM et al. Containing COVID-19 Among 627,386 Persons in Contact With the Diamond Princess Cruise Ship Passengers Who Disembarked in Taiwan: Big Data Analytics. J Med Internet Res.2020 May 5;22(5):e19540.

15 Strategy to prevent Covid-19 community transmission. CDC Taiwan, April 6, 2020 (in mandarin, translation available upon request) https://www.cdc.gov.tw/Uploads/6f836467-3f73-4153-86fb-eb96a1acf437.pdf

16 Infectious control guideline for medical institutions in response to COVID-19, CDC Taiwan. June 2^nd^ 2020 2020 (in mandarin, translation available upon request)

17 M-Y Yen et al. Taiwan’s Traffic Control Bundle and the Elimination of Nosocomial Severe Acute Respiratory Syndrome Among Healthcare Workers, J Hosp Infect. 2011 Apr;77(4):332–7.

18 Contingency recommendations for hospitals in response to COVID-19 (Wuhan pneumonia) confirmed cases. CDC Taiwan. May 5^th^ 2020 (https://fightcovid.edu.tw/cdc-guidelines/contingency-recommendation)

19 Established according to the Regulations Governing Operation of the Communicable Disease https://law.moj.gov.tw/ENG/LawClass/LawAll.aspx?pcode=L0050014

20 Infectious control guideline for Long-Term Care Organizations in Response to COVID-19, CDC Taiwan, February 22^nd^ (in mandarin, translation available upon request)

21 Guideline for contingency plan for Health and Welfare Institution (boarding homes) in response to COVID- 19, CDC Taiwan. May 20^th^ 2020 (in mandarin, translation available upon request)

22 Criteria for school suspension in response to Covid-19. Ministry of education, February 12^th^, 2020 (in mandarin, translation available upon request)

23 Taiwan Social Distancing APP https://covirus.cc/social-distancing-app-intro.html

24 Special Act for Prevention, Relief and Revitalization Measures for Severe Pneumonia with Novel Pathogens https://law.moj.gov.tw/ENG/LawClass/LawAll.aspx?pcode=L0050039

25 open source App provided by the Digital Minister Tang: - Frontend https://github.com/gdg-twhk/mask-web - iOS/Android app https://github.com/gdg-twhk/mask-app - Backend API https://github.com/gdg-twhk/mask-gae App to support Korea: https://github.com/kiang/covid19-kr-masks/

26 Special Act for Prevention, Relief and Revitalization Measures for Severe Pneumonia with Novel Pathogens https://law.moj.gov.tw/ENG/LawClass/LawAll.aspx?pcode=L0050039

27 Open and transparent information. Ministry of Health and Welfare, https://covid19.mohw.gov.tw/en/cp-4772-53699-206.html

